# Burden of rodent-borne viruses in rodents and zoonotic risk in human in Cambodia: a descriptive and observational study

**DOI:** 10.1101/2025.02.09.25321973

**Authors:** Julia Guillebaud, Janin Nouhin, Vibol Hul, Thavry Hoem, Oudamdaniel Yanneth, Mala Sim, Limmey Khun, Y Phalla, Sreymom Ken, Leakhena Pum, Reaksa Lim, Channa Meng, Kimtuo Chhel, Sithun Nuon, Sreyleak Hoem, Kunthy Nguon, Malen Chan, Sowath Ly, Erik A. Karlsson, Jean-Marc Reynes, Anavaj Sakunthabhai, Philippe Dussart, Veasna Duong

## Abstract

**Background:** Rodent-borne viruses, including hantaviruses, arenaviruses, and rodent hepatitis virus (HEV-C), pose significant health threats to humans, causing severe diseases such as hepatitis, respiratory illness, and hemorrhagic fevers. In Cambodia, data on these viruses remain limited, and their burdens on human health are unknown. This study investigated the presences of these viruses in rodents and assessed potential human exposure across diverse environmental and socio-economic contexts in Cambodia.

**Methods:** The study was conducted in urban, semi-urban, and rural areas of Cambodia during the rainy (2020) and dry seasons (2022). Rodents were screened for arenavirus, hantavirus, and HEV-C using RT-PCR. Human serum samples from the same site were tested for IgG antibodies using ELISA. Factors associated with virus spillover into humans were analyzed.

**Findings:** Among 750 rodents, 9.7% carried at least one virus: 5.2% arenavirus, 3.3% hantavirus, and 1.9% HEV-C. Infection rates were highest in urban (14.5%), followed by semi-urban (11.9%) and rural (2.1%) interfaces. Arenavirus was more prevalent during the rainy season, while hantavirus and HEV-C remained consistent across seasons. Seroprevalence in human was 12.7% for arenavirus, 10.0% for hantavirus, and 24.2% for HEV. Higher arenavirus seroprevalence was associated with urban recidency and lower education level. Hantavirus seroprevalence was associated with urban residency, acute hepatitis history, and flood-prone living areas. HEV seroprevalence increased with urban residency, increasing age, and medical condition history.

**Interpretation:** Our findings highlighted the need for rodent control, improved market infrastructure, enhanced waste management, and public awareness on hygiene practices and zoonotic risks, especially in urban and high-risk areas.

## INTRODUCTION

The emergence of zoonotic viruses is driven by a complex interplay of environmental, ecological, and socio-economic factors. Deforestation, agricultural expansion, urbanization, and encroachment of human settlements disrupt animal habitats, alter behavior, and intensify human-animal interactions, increasing the likelihood of spillover.^1,2^ Once a spillover occurs, human-to-human transmission can be amplified by globalization and modern transportation infrastructure.^3^ Understanding the dynamics of virus spillover, from animal reservoirs to human populations, is crucial for developing effective public health surveillance and mitigation strategies.

Cambodia, a Least Developed Country in the Greater Mekong Subregion of Southeast Asia, harbors diverse ecosystems with high biodiversity including tropical rainforests, wetlands, and rice fields. Rapid economic transformation over the past two decades has led to increased urbanization and significant environmental changes.^4^ Urban expansion into biodiverse ecosystems has heightened the potential for zoonotic spillover events. However, the understanding of zoonotic virus prevalence and their associated risks remains relatively limited. Investigating how changes in the built environment and encroachment can impact zoonotic risk is vital for anticipating and mitigating potential outbreaks.

Several rodent-borne pathogens have been detected in Cambodia, including hantaviruses and arenaviruses.^5,6^ Hantaviruses have been documented in rodents since 1998, with reports from the capital city, Phnom Penh, as well as the southern and eastern provinces of the country.^5^ A relatively high proportion of recent hantavirus infections in humans was reported, with 71 out of 459 individuals (15.5%) had IgG four-fold rise in titer or seroconversion.^7^ Recently, the Cardamon variant of Wenzhou virus has been identified in brown rats (*Rattus norvegicus*) and Pacific rats (*R. exulans*). Infection in human has been reported in febrile cases with respiratory symptoms.^6^ Information on rodent-borne Hepatitis E virus (HEV-C) is limited in Cambodia. A virome analysis of rodent lungs collected from 2006 to 2018 found HEV-C partial genome sequences.^8^ HEV-C has also been in neighboring countries, including Thailand, Laos PDR, and Vietnam, as well China, Indonesia, and Japan.^8–12^

These three rodent-borne viruses represent significant public health threats due to their potential to cause severe disease, ranging from hepatitis to respiratory and hemorrhagic fevers.^13–16^ Transmission to humans typically occurs through direct or indirect contact with infected animals, contaminated excreta, or environment.^15–17^ Despite these risks, comprehensive assessment of rodent-borne pathogens across diverse human-animal interfaces in Cambodia are rare.

This study aimed to address this knowledge gap by investigating the presence of rodent-borne viruses— arenaviruses, hantaviruses, and HEV-C—in small mammals and assessing evidence of human exposure across different environmental and socio-economic contexts in Cambodia. Investigation focused on the capital city, where high rodent-human interaction is anticipated, leading to an elevated risk of transmission. Subsequently, we extended our study to include interfaces representing Cambodia’s diverse landscapes, including semi-urban and rural areas.

## MATERIALS AND METHODS

### Ethics approvals

Study protocols were approved by the Cambodian National Ethics Committee for Health Research (NECHR) (N° 320 NECHR, N° 132 NECHR, N° 215 NECHR, and N° 256 NECHR). Written informed consent for the use of demographic and biological data was obtained from adult participants or guardians of participants under 18 years old. Animals capture and handling followed American Society of Mammalogists guidelines.^18,19^ Statutory study permission was granted by the Forestry Administration of the Cambodian Ministry of Agriculture, Forestry and Fisheries, the national authority responsible for wildlife research.

### Study timing and locations

This descriptive study was conducted across three distinct interfaces in Cambodia—urban, semi-urban, and rural—during the 2020 rainy season and 2022 dry season (Figure 1A). Study sites were selected to represent an urbanization gradient, reflecting different levels of human development and interaction with the environment. Phnom Penh, the capital city, was classified as the urban interface due to its dense population of approximately 2.1 million inhabitants. Five major markets—Phsar Chas, Phsar Kandal, Orussey, Central and Chbar Ampov—were chosen based on their size and location, prioritizing sites with high likelihood of rodent-human interactions and the potential for the widespread distribution contamination within the surrounding neighborhoods. Preah Sihanouk province, located on the western coast, was selected as a semi-urban interface, representing a region undergoing rapid urbanization, transitioning from the original hills and rice fields to a more developed landscape. Three villages—Veal Thum, Veal Meas, and Boeng Vaeng—were selected. Kampong Cham province, known for its predominantly agrarian landscape and extensive rice fields, served as the rural interface, with the villages of Roung Kou, Krasang Pul, and Toul Ampil selected.

**Figure 1:**
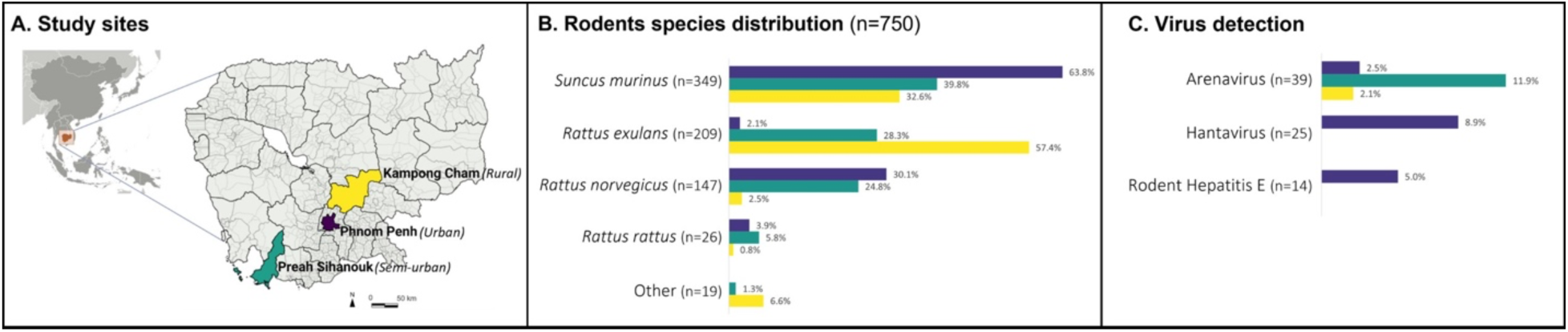
Study sites, rodent species, and virus prevalence in rodents. (A) Study site: Trapping was conducted twice at each site. Session 1 took place between July and September 2020 across all interfaces. Session 2 occurred between January and February 2021 in urban (Phnom Penh, depicted in purple) and rural (Kampong Cham province, depicted in yellow) interfaces, and in January 2022 in Semi-urban (Sihanoukville province, depicted in green). (B) Distribution of rodent species: The chart illustrates the overall distribution of rodent species identified in each interface. “Other” category includes: Bandicota savilei (n=1), Rattus argentiventer (n=2), Mus sp. (n=9), Rattus sp. (n=7). (C) Virus detection in rodents: The presences of arenavirus and hantavirus was screened using pooled organ samples (kidney, spleen, liver and lung) from individual rodent. HEV-C was screened from individual rodent liver samples.

### Sample and data Collection

At each site, samples were collected from rodents as well as humans residing or working in the same areas. Initial sample and data collections occurred from July to September 2020, with a planned follow-up 6 months later. However, travel restrictions and other non-pharmaceutical interventions in response to the coronavirus disease 2019 (COVID-19) pandemic delayed the second sampling until March—April 2022.

### Rodent sample collection

Rodents were live-trapped using locally made traps placed along pathways known to be frequented by rodents. At each site, 50 to 60 traps were deployed each evening for two to three consecutive nights, to capture at least 30 small mammals per session. In the morning, trapped rodents were humanely euthanized following the American Veterinary Medical Association Guidelines.^19^ Key morphological parameters, including sex, body length, weight, reproductive status, approximate age (juvenile vs. adult), were recorded for species identification. Lung, liver, spleen, and kidney tissues were collected and stored individually in 1 mL of Virus Transport Medium prepared as previously described.^20^ All samples were preserved on-site in liquid nitrogen and subsequently transferred to the Virology Unit, Institut Pasteur du Cambodge (IPC), where they were stored at −80°C.

### Human sample and data collection

Market workers (≥18 years) from urban interface and villagers (≥2 years) from semi-urban and rural interfaces, residing near rodent trapping sites, were invited to participate in the study. Each participant was requested to provide one blood sample. After obtaining informed consent, trained interviewers conducted individual interviews in Khmer to collect basic socio-epidemiological data and information on potential exposure factors using paper-based questionnaires. Venous blood (5 mL) was collected in a dry tube and temporarily stored at +4°C in a cool box, and transported daily to IPC’s Virology Unit. Blood samples were centrifuged (10 min, 2,000 rpm). Serum was aliquoted and stored at −80°C until analysis.

### Rodent-borne virus detection in animal samples

Approximately 10 mg (equivalent to 3 mm^3^) of liver and pooled organs (kidney, spleen, liver, and lung) of each rodent were homogenized using MagNA Lyzer Instrument and bead system (Roche, Basel, Switzerland). Viral RNA was extracted from 200 μL of the supernatant of the homogenized tissue using Direct-Zol RNA MiniPrep kits (Zymo Research) according to manufacturer’s instructions.

Rodent-borne HEV-C screening was performed on RNA extracted from liver samples using a differntial duplex one-step real-time RT-PCR to distinguish it from and human-associated hepatitis E virus (HEV-A).^21,22^ Detection of hantavirus and arenavirus was conducted on RNA extracted from pooled organs using RT-PCR targeting the 412 bp and 395 bp L segment, respectively, as previously described.^23,24^

### Rodent speciation

Speciation was performed for all rodents positive for at least one pathogen of interest, as well as five negative rodents of each taxon, using Sanger sequencing of vertebrate mitochondrial *cytochrome oxidase subunit 1 (CO1)* and *cytochrome b (Cyt b)* DNA as previously described.^25^ Sequence chromatograms were reviewed through visual inspection using CLC Main Workbench software (CLC Bio). Nucleotide sequences were analyzed by NCBI BLAST to determine sequence identity.^26^

### *CO1* and *Cyt b* nucleotide sequence accession number

*CO1* and *Cyt b* sequences were submitted to GeneBank and registered under accession numbers: XXXXXXXX – XXXXXXXX and XXXXXXXX – XXXXXXXX, respectively

### IgG antibody detection in human serum

We tested participant sera for IgG antibodies against the three viruses of interest. Anti-HEV IgG antibodies were screened using a commercial HEV IgG enzyme-linked immunosorbent assay (ELISA) kit (Beijing Wantai Biological Pharmacy Enterprise, Beijing, China) according to manufacturer’s instructions. Anti-arenavirus and anti-hantavirus IgG antibodies using an in-house ELISA.^6,27^ Positive samples identified during the primary screening were repeated. Presence of IgG antibodies was considered previous exposure to the pathogen. Seroconversion toward a virus was defined as the transition from IgG seronegativity at baseline to seropositivity in the follow-up visit.

### Statistical analysis

Descriptive statistics (Chi-square test, Fisher’s exact test) were used to assess the relationships between categorical variables. Exposure to arenavirus, hantavirus, and HEV was defined by a seropositive result (positive IgG). Each serological status was analyzed using a generalized linear model with multivariate logistic regression to explore the associations between viral seroprevalence and explanatory variables such as sociodemographic characteristics and potential risk factors. In the final model selection process, while we primarily relied on the stepwise model selection to identify the most parsimonious model based on the Akaike Information Criterion (AIC), we also forced the inclusion of specific variables (e.g., age, sex, and interface) into each model. These variables were retained due to their known biological relevance or potential role as confounders, even though they did not meet the AIC-based selection criteria. A p-value of 0.05 was considered statistically significant. Statistical analysis was performed using R software.^28^

## RESULTS

### Detection of Arenavirus, Hantavirus, and HEV-C in rodents

#### Distribution of rodent species by interface

A total of 750 rodents were captured in the two sampling sessions across the three defined interfaces (Figure 1A). Of these, 282 (37.6%) were from the urban interface, with 114 (40.4%) collected during the rainy season and 168 (59.6%) during the dry season. The semi-urban interface accounted for 226 rodents (30.1%), with 137 (60.6%) collected in the rainy season and 89 (39.4%) in the dry season. In the rural interface, 242 rodents (32.3%) were captured, with 85 (35.1%) collected during the rainy season and 157 (64.9%) in the dry season.

Across all interfaces, the most frequently captured species was *Suncus murinus* (Asian house shrew: 46.5%; 349/750), followed by *Rattus exulans* (Pacific rat: 27.9%; 209/750), *Rattus norvegicus* (Brown rat: 19.6%; 147/750) and *Rattus rattus* (Black rat: 3.5%; 26/750). Less common species (<2% each) were grouped as “Other” (2.5%; 19/750) and included *Mus sp.* (unidentified mouse species: 1.2%; 9/750), and *Rattus sp.* (unidentified rat species: 0.9%; 7/750), *Rattus argentiventer* (rice field rat: 0.3%; 2/750), and *Bandicota savilei* (Savile’s bandicoot rat: 0.1%; 1/750).

Rodent specie distribution varied significantly across the interfaces (p<0.001) (Figure 1B). *S. murinus* dominated the urban interface (63.8%; 180/282), followed by *R. norvegicus* (30.1%; 85/282). The semi-urban interface showed a more balanced distribution, with *S. murinus*, *R. exulans*, and *R. norvegicus* accounting for 39.8% (90/226), 28.3% (64/226), and 24.7% (56/226) of collected rodents, respectively. In the rural interface, *R. exulans* was the predominant species (57.4%, 139/242), followed by *S. murinus* (32.6%, 79/242).

#### Virus prevalence across Interfaces

Of the 750 collected rodents, 73 (9.7%) carried at least one virus of interest, with 68 (9.1%) infected with one virus and 5 (0.6%) co-infected with two viruses. Specifically, 39 rodents (5.2%) were positive for arenavirus, 25 (3.3%) for hantavirus, and 14 (1.9%) for HEV-C. The proportion of infected rodents with at least one virus significantly varied among interfaces, with the urban interface displaying the highest proportion of positive animals (14.5%, 41/282), followed by the semi-urban (11.9%, 27/226) and rural (2.1%, 5/242) interfaces (p<0.001). Arenavirus was detected in all three interfaces, with prevalences of 2.5% (7/282) in urban, 11.9% (27/226) in semi-urban, and 2.1% (5/242) in rural areas (p<0.001). Hantavirus and HEV-C were exclusively detected in the urban interface, with prevalences of 8.9% (25/282) and 5.0% (14/282), respectively (Figure 1C). All of five rodents with co-infections were from urban interface. These included four animals with HEV-C/hantavirus co-infection and one animal with hantavirus/arenavirus co-infection.

#### Seasonal variations in virus prevalence

The rate of infection for at least one virus was significantly higher during the rainy season (12.8%, 43/336) versus the dry season (7.2%, 30/414) (p=0.01). The arenavirus prevalence was significantly higher during the rainy season, particularly in the semi-urban interface (overall: 7.7% in the rainy season vs. 3.1% in the dry season, p=0.005; semi-urban: 16.8% in the rainy season vs. 4.5% in the dry season, p=0.006). Detection rates for hantavirus and HEV-C were consistent across seasons (Table 1).

**Table 1:** Detection rate of arenavirus, hantavirus, and HEV-C in rodents per interface and season.

|  | Overall |  |  | Urban |  |  | Semi-urban |  |  | Rural |  |  |
| --- | --- | --- | --- | --- | --- | --- | --- | --- | --- | --- | --- | --- |
|  | Rainy season<br>(n=336) | Dry season<br>(n=414) | p value* | Rainy season<br>(n=114) | Dry season<br>(n=168) | p value* | Rainy season<br>(n=137) | Dry season<br>(n=89) | p value* | Rainy season<br>(n=85) | Dry season<br>(n=157) | p value* |
| Arenavirus | 26 (7.7%) | 13 (3.1%) | 0.005 | 3 (2.6%) | 4 (2.4%) | 1.000 | 23 (16.8%) | 4 (4.5%) | 0.006 | 0 (0%) | 5 (3.2%) | 0.165 |
| Hantavirus | 13 (3.9%) | 12 (2.9%) | 0.46 | 13 (11.4%) | 12 (7.1%) | 0.217 | 0 (0%) | 0 (0%) | .. | 0 (0%) | 0 (0%) | .. |
| HEV-C | 8 (2.4%) | 6 (1.4%) | 0.35 | 8 (7.0%) | 6 (3.6%) | 0.219 | 0 (0%) | 0 (0%) | .. | 0 (0%) | 0 (0%) | .. |

Data are n (%), \*p value was calculated using a Chi-squared or Fisher's exact tests

#### Variations in host species

Among the 39 rodents infected with arenavirus, the majority were *R. exulans* (74.4%; 29/39), followed by *R. norvegicus* (17.9%; 7/39) and *R. rattus* (7.7%; 3/39). Of the 25 rodents infected with hantavirus, 92% (23/25) were *R. norvegicus*, with one case each in *R. rattus* (4%) and *S. murinus* (4%). All 14 rodents infected with HEV-C were *R. norvegicus*. All 4 rodents co-infected with HEV-C/hantavirus were *R. norvegicus*, while the rodent co-infected with hantavirus/arenavirus was *R. rattus*.

### Evidence of spillover of rodent-borne viruses in humans

During the initial visit (July—September 2020), 788 human participants were enrolled across the three interfaces. Of these, 304 (38.6%) participants were from urban areas, 288 (36.5%) from semi-urban areas, and 196 (24.9%) from rural areas. Females accounted for 64.5% (n=508) of the participants, with a median age of 38 years (interquartile range, IQR: 13-54). Table 2 provides a summary of sociodemographic characteristics and potential risk factors associated with each interface type.

**Table 2:** Baseline characteristic.

|  | Overall | Urban | Semi-urban | Rural | p value* |
| --- | --- | --- | --- | --- | --- |
|  | (n=788) | (n=304) | (n=288) | (n=196) |  |
| Gender |  |  |  |  | 0.006 |
| Female | 508 (64.5%) | 213 (70.1%) | 166 (57.6%) | 129 (65.8%) | .. |
| Male | 280 (35.5%) | 91 (29.9%) | 122 (42.4%) | 67 (34.2%) | .. |
| Age, year | 38 (13-54) | 50 (40-60) | 17 (10-47) | 15 (9-41) | <0.001 |
| Age group, year |  |  |  |  | <0.001 |
| <18 | 255 (32.4%) | 105 (53.6%) | 149 (51.7%) | 1 (0.3%) | .. |
| 18-35 | 112 (14.2%) | 27 (13.8%) | 33 (11.5%) | 52 (17.1%) | .. |
| 36-50 | 174 (22.1%) | 27 (13.8%) | 46 (16.0%) | 101 (33.2%) | .. |
| ≥50 | 247 (31.3%) | 37 (18.9%) | 60 (20.8%) | 150 (49.3%) | .. |
| Education |  |  |  |  | <0.001 |
| No school | 127 (16.1%) | 48 (15.8%) | 48 (16.7%) | 31 (15.8%) | .. |
| Primary | 405 (51.4%) | 129 (42.4%) | 156 (54.2%) | 120 (61.2%) | .. |
| Secondary and higher | 256 (32.5%) | 127 (41.8%) | 84 (29.2%) | 45 (23.0%) | .. |
| Occupation |  |  |  |  | <0.001 |
| Agricultural worker | 42 (5.3%) | 0 (0.0%) | 6 (2.1%) | 36 (18.4%) | .. |
| Seller | 311 (39.5%) | 245 (80.6%) | 49 (17.0%) | 17 (8.7%) | .. |
| Other | 435 (55.2%) | 59 (19.4%) | 233 (80.9%) | 143 (73.0%) | .. |
| Residency |  |  |  |  |  |
| Less than 1 year | 14 (1.8%) | 4 (1.3%) | 3 (1.0%) | 7 (3.6%) | .. |
| More than 1 year | 212 (26.9%) | 146 (48.0%) | 29 (10.1%) | 37 (18.9%) | .. |
| Entire life | 562 (71.3%) | 154 (50.7%) | 256 (88.9%) | 152 (77.6%) | .. |
| Any medical condition |  |  |  |  | .. |
| Previous | 146 (18.5%) | 84 (27.6%) | 39 (13.5%) | 23 (11.7%) | <0.001 |
| Chronic | 49 (6.2%) | 35 (11.5%) | 7 (2.4%) | 7 (3.6%) | <0.001 |
| Alcohol consumption | 599 (76.0%) | 198 (65.1%) | 251 (87.2%) | 150 (76.5%) | <0.001 |
| Any acute hepatitis history | 15 (1.9%) | 6 (2.0%) | 9 (3.1%) | 0 (0.0%) | 0.03 |
| Living conditions |  |  |  |  | .. |
| Flooded area | 118 (15.0%) | 19 (6.2%) | 94 (32.6%) | 5 (2.6%) | <0.001 |
| Swine farm nearby | 116 (14.7%) | 44 (14.5%) | 30 (10.4%) | 42 (21.4%) | 0.004 |
| Slaughter house nearby | 66 (8.4%) | 58 (19.1%) | 7 (2.4%) | 1 (0.5%) | <0.001 |
| Any wild animal hunting experience | 87 (11.0%) | 15 (4.9%) | 52 (18.1%) | 20 (10.2%) | <0.001 |
| Dietary habits |  |  |  |  | .. |
| Raw pork meat | 64 (8.1%) | 21 (6.9%) | 34 (11.8%) | 9 (4.6%) | 0.011 |
| Pork sausage | 148 (18.8%) | 50 (16.4%) | 63 (21.9%) | 35 (17.9%) | 0.20 |
| Raw shells | 311 (39.5%) | 123 (40.5%) | 102 (35.4%) | 86 (43.9%) | 0.20 |
| Rodent meat | 13 (1.6%) | 5 (1.6%) | 4 (1.4%) | 4 (2.0%) | 0.88 |
| Experience of direct contacts with pork | 514 (65.2%) | 229 (75.3%) | 162 (56.2%) | 123 (62.8%) | <0.001 |
| Experience of direct contacts with poultry | 502 (63.7%) | 221 (72.7%) | 163 (56.6%) | 118 (60.2%) | <0.001 |
| Experience of contacts with rodents |  |  |  |  | .. |
| Indirect contacts | 721 (91.5%) | 296 (97.4%) | 252 (87.5%) | 173 (88.3%) | >0.9 |
| Direct contacts | 135 (17.1%) | 53 (17.4%) | 58 (20.1%) | 24 (12.2%) | 0.076 |
| Got bitten by a rodent | 45 (5.7%) | 28 (9.2%) | 11 (3.8%) | 6 (3.1%) | 0.003 |

Data are n (%), median (IQR), \*p value was calculated using a Chi-squared test

Exposure to rodent-borne viruses was identified in a proportion of participants, with seroprevalence rates of 12.7% (n=100) for arenavirus, 10.0% (n=79) for hantavirus, and 24.2% (n=191) for HEV among the total cohort. Analysis by interface revealed notable differences in seroprevalence, with urban areas exhibiting the highest rates compared to semi-urban and rural areas. For arenavirus, 21.7% (66/304) of individuals were seropositive in urban areas, compared to 5.6% and 9.2% in semi-urban and rural areas, respectively (p<0.001). For hantavirus, 13.2% (40/304) were positive in urban areas, compared to 9.4% and 6.1% in semi-urban and rural areas, respectively (p=0.03). For HEV, 41.1% (125/304) were positive in urban areas, compared to 14.9% and 11.7% in semi-urban and rural areas, respectively (p<0.001).

### Factors associated with rodent-borne virus spillover into humans

Multivariate analysis identified several factors significantly associated with higher seroprevalence of rodent-borne viruses in humans (Figure 2). Arenavirus IgG seroprevalence was associated with urban residency (adjusted odds ratio [ORa]=2.5, 95% CI: 1.3–4.9, p=0.01) and lower education (ORa=2.2, 95% CI: 1.1–4.8, p=0.03). Hantavirus IgG seroprevalence was associated with urban residency (ORa=2.3, 95% CI: 1.1–5.3, p=0.03), a history of acute hepatitis (ORa=5.3, 95% CI: 1.5–16.3, p=0.01), and living in flooding-prone areas (ORa=2.0, 95% CI: 1.0–3.9, p=0.04). HEV IgG seroprevalence was linked to urban residency (ORa=2.0, 95% CI: 1.1–3.6, p=0.02), increasing age (18–35 years: ORa=9.3, 95% CI: 2.7–43.3, p<0.001; 36–50 years: ORa=37.6, 95% CI: 12.3–164, p<0.001; ≥50 years: ORa=75.9, 95% CI: 25.5–329, p<0.001), and a history of medical conditions (ORa=1.6, 95% CI: 1.0–2.4, p=0.04). Interestingly, a higher education level (above primary school) was associated with lower HEV IgG seroprevalence (ORa=0.5, 95% CI: 0.3–0.9, p=0.04).

**Figure 2:**
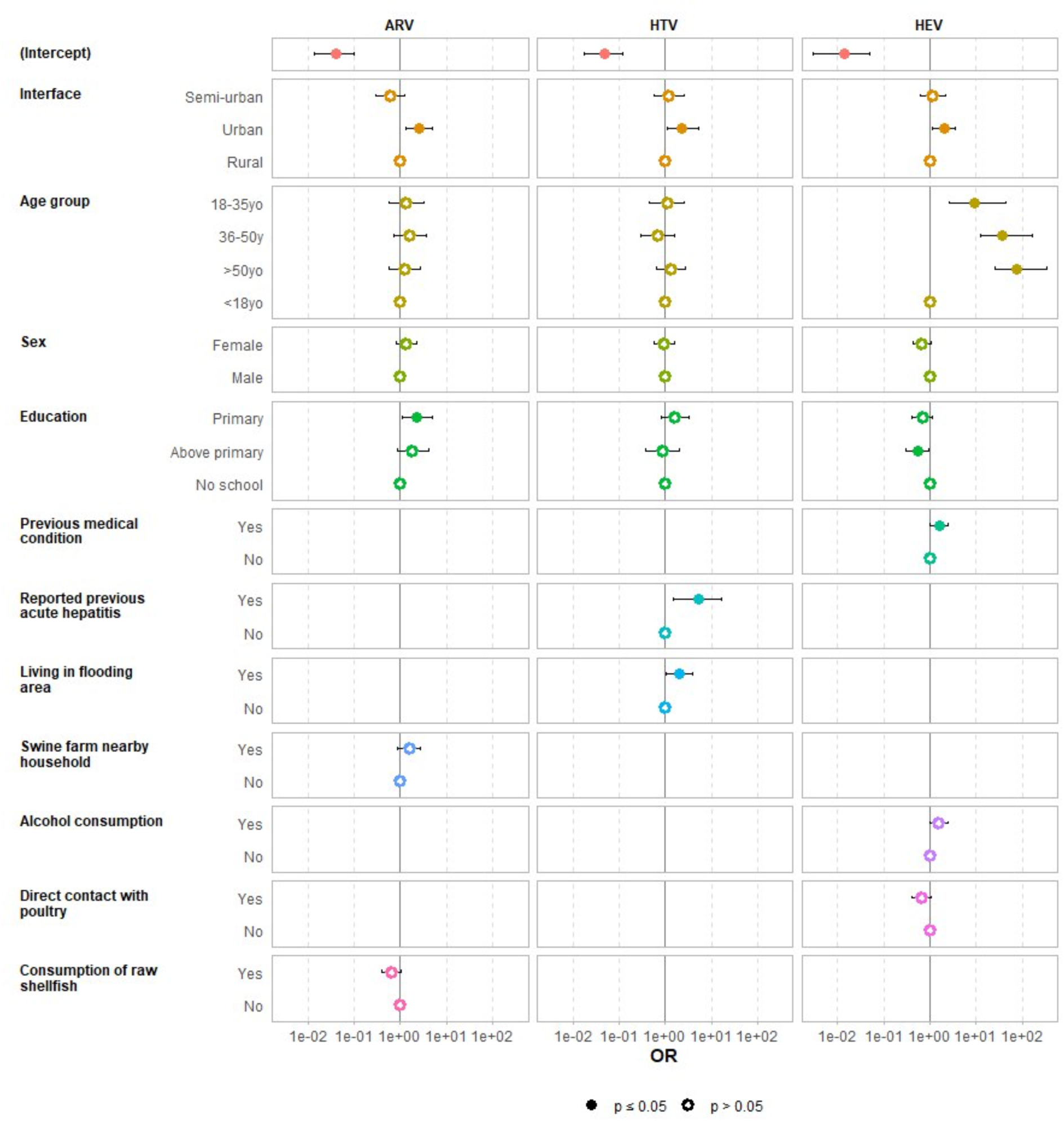
Forest plot showing factors associated with exposure to arenavirus (ARV), hantavirus (HTV), and hepatitis E virus (HEV) in human population. OR: odds ratio; CI: confidence interval

### Follow-up visits and seroconversion rates

In 2022, 555 (70.4%) of 788 included participants were able to participate in the follow-up visit. Of these, 39.1% (217/555) participants were from urban, 36.6% (203/555) from semi-urban, and 24.3% (135/555) from rural interfaces (Table 3). Among participants who were seropositive at inclusion, 36.4% (28/77) maintained their seropositive status for anti-arenavirus IgG, 89.5% (51/57) for anti-hantavirus IgG, and 96.2% (125/130) for anti-HEV IgG.

**Table 3:** Follow-up serological results of participants for each tested virus and per interface.

|  | Overall | Urban | Semi-urban | Rural |
| --- | --- | --- | --- | --- |
| **Inclusion, n (%)** | **788 (100%)** | **304 (38.6%)** | **288 (36.5%)** | **196 (24.9%)** |
| **Follow-up, n (%)** | **555 (70.4%)** | **217 (71.4%)** | **203 (70.5%)** | **135 (68.9%)** |
| **Arenavirus** |  |  |  |  |
| IgG seropositive at inclusion | 77 (13.9%) | 52 (24.0%) | 11 (5.4%) | 14 (10.4%) |
| Participants remains seropositive | 28 (36.4%) | 11 (21.2%) | 6 (54.5%) | 11 (78.6%) |
| IgG seroconversion* | 84 (17.6%) | 10 (6.1%) | 21 (10.9%) | 53 (43.8%) |
| **Hantavirus** |  |  |  |  |
| IgG seropositive at inclusion | 57 (10.3%) | 29 (13.4%) | 18 (8.9%) | 10 (7.4%) |
| Remain seropositive | 51 (89.5%) | 27 (93.1%) | 17 (94.4%) | 7 (70.0%) |
| IgG seroconversion* | 6 (1.2%) | 2 (1.1%) | 1 (0.5%) | 3 (2.4%) |
| **HEV** |  |  |  |  |
| IgG seropositive at inclusion | 130 (23.4%) | 85 (39.2%) | 32 (15.8%) | 13 (9.6%) |
| Remain seropositive | 125 (96.2%) | 82 (96.5%) | 31 (96.9%) | 12 (92.3%) |
| IgG seroconversion* | 9 (2.1%) | 6 (4.5%) | 2 (1.2%) | 1 (0.8%) |

Data are n (%), \*Participant presenting a seronegative result at inclusion and seropositive result at follow-up visit

Seroconversion was observed in 17.6% (84/478) of participants for arenavirus, 1.2% (6/498) for hantavirus, and 2.1% (9/425) for HEV. For arenavirus, the majority of seroconversions occurred in participants from rural interfaces (63.1%), followed by semi-urban (25.0%) and urban (11.9%) interfaces (p<0.001). Participants who had lived in the study area their entire lives showed a significantly higher seroconversion rate (72.6%, 61/84) than those residing there for less than a year (4.8%, 4/84) or more than a year (22.6%, 19/84) (p=0.02).

## DISCUSSION

Rodent-borne viruses, including arenavirus, hantavirus, and HEV-C, pose a significant public health concern in Cambodia. The country’s tropical climate, widespread agricultural activities, and rapid urbanization expansion create ideal conditions for rodent population growth and increased human-rodent interactions, heightening the risk of zoonotic spillover.^29^ Inadequate housing structures and limited pest control further amplifies exposure risks. Additionally, gaps in healthcare infrastructure and limited awareness of these pathogens contributes to their neglect. Understanding the prevalence and distribution of these viruses is essential for designing targeted interventions to mitigate their impact. This study explored the prevalence of these viruses in rodents and humans across urban, semi-urban, and rural interfaces during both rainy and dry seasons.

In line with previous studies in Vietnam, Indonesia, and China, all three viruses of interest were detected in Cambodia, with the highest prevalence observed in urban rodents (2.5% for arenavirus, 8.9% for hantavirus, and 5.0% for HEV-C).^11,30–32^ Arenavirus showed higher detection rates in the rainy season, while no seasonal trend was observed for hantavirus and HEV-C. The presence of multiple rodent-borne viruses in urban rodents can be attributted to densely populated rodent communities, which facilitate virus transmission. Our urban rodents were captured from markets, where abundant food, poor waste management, and lack of rodent control programs create favorable conditions for rodent population growth and interspecies virus transmission. Additionally, those rodents live in the sewage system which often flooded during rainy season affecting their habitat and forcing them to live densely. In contrast, rodents in semi-urban and rural interfaces were mainly collected from households not affected by flooding.

The seroprevalences among humans were 12.7% for arenavirus, 10.0% for hantavirus, and 24.2% for HEV at baseline. A clear gradient was observed, with urban setting showing the highest rate (22% for arenavirus, 13% for hantavirus, and 41% for HEV) compared to rural areas (9%, 6%, and 12%, respectively). The hantavirus seroprevalence in humans aligns with previous studies in Cambodia.^7^ Lower seroprevalence rates have been reported in Vietnam in various populations: 1.1% of healthy donors and 2.3% of febrile patients in North Vietnam in 2009 (33), and 3.7% of people living in farming communities the Mekong Delta in 2015.^34^ The seroprevalence of HEV found in our study does not reflect the actual HEV-C seroprevalence, since we used the commercial Wantai ELISA kit which does not differentiate HEV-A and HEV-C.^35^ Nevertheless, this kit has been shown to cross-react with antibodies against both HEV-A and HEV-C.^36^ This underscores the urgent need to develop sensitive diagnostic assays specific to HEV-C for human serological investigations.

Urban residency was strongly associated with elevated risk of virus exposure, consistent with the high virus prevalence in urban rodents. However, arenavirus seroprevalence was unexpectedly high in urban human population despite low circulation in rodents. One explanation could be the low density of competent hosts in this area, as most arenavirus infected rodents were *Rattus exulans* which commonly found in both semi-urban and rural interfaces. Additionally, it is possible that a single infected rodent could rapidly transmit the virus to multiple humans due to the high rodent-human interactions in densely urban market setting.

Interestingly, history of acute hepatitis was significantly associated with hantavirus seropositivity. This association has been reported in a few cases in Malysia and Japan. One hypothesis suggests that hantavirus infection may trigger acute exacerbation of autoimmune liver disease. However, community-acquired hepatitis could not be excluded. Although hantavirus infections rarely cause hepatitis without renal manifestation, this warrants further investigation.^37,38^ No instances of renal manifestation were reported in our study. Additionally, living in flood-prone areas was associated with higher hantavirus seroprevalence, likely due to rodent habitat disruption and environmental contamination caused by flooding.^39^ For HEV, older age was a significant risk factor, as described in previous studies, reflecting cumulative lifetime exposure, while higher education levels were protective, likely due to better health literacy and hygiene practices.^40,41^

Several limitations to this study should be noted. First, the delay in follow-up due to the COVID-19 pandemic extended the timeframe between visits, preventing evaluation of short-term exposure incidence. Second, seroprevalence estimates may be overestimated due to potential cross-reactivity with other closely related viruses and could require further confirmatory testing. Third, the ELISA kit used for HEV-C serological assay cannot differentiate HEV-A and HEV-C, reducing the specificity in serological findings. Further studies will focus developing and employing for more robust diagnostic tools and methodologies to reduce these issues.

Our findings underscore the critical need for effective rodent control measures, improved market infrastructure, and enhanced waste management to reduce the risk of rodent-borne virus transmission to humans. Awareness campaigns promoting hygiene and education on zoonotic risks are also essential, particularly in urban and high-risk areas. The evidence of virus exposure in humans highlights a clear need for continued, active, longitudinal surveillance and diagnostic advancement to monitor rodent-borne virus dynamics and assess their clinical impact on human health at a countrywide level. Furthermore, these results inform medical professionals regarding diseases caused by rodent-borne viruses, enabling better differential diagnoses and improved patient cares. Indeed, this study underscores the value of a One

Health approach in understanding the complex interplay between environmental, social, and biological factors that drive zoonotic virus transmission. The findings serve as a reminder that addressing zoonotic pathogens is not solely about mitigating risks—it is about fostering sustainable human-animal coexistence in shared ecosystems.

## CONTRIBUTORS

VH, JG, JN, PD, and VD conceived and designed the study. JG, TH, OY, and MS performed field work to collect samples. JG, VH and SL prepared questionnaire for human epidemiological data. KN, and MC collected human epidemiological data. OY, MS, LK, LP, RL, KC, SN, and SH carried out molecular assays for virus screening and detection. OY, MS, PY and SK performed serological assays. JG, VH, TH, and JN assessed and verified all data for analysis in this study. JG and OY performed statistical analysis. J-MR led technology transfer of serological assay to detect the exposure of hantavirus. JG and JN drafted the manuscript. EAK, VD and AS provided critical revision of the manuscript and intellectual inputs. All authors reviewed and approved the final draft for submission. All authors had full access to all the data in the study and had final responsibility for the decision to submit for publication.

## DATA SHARING

All data included in the study are available from JG and JN upon request.

## ACKNOWLEDGEMENT

We are thankful to all relevant authorities from the Ministry of Agriculture, Forestry and Fisheries of Cambodia for their support in facilitating this work. We particularly acknowledge the Department of Wildlife and Biodiversity under the Forestry Administration for their continuous support and participation during field data collection. We are grateful to local authorities for facilitating all field work in selected areas and provide valuable human resources. We especially express our gratitude to all participants from this study. We are grateful to R. Johne (Federal Institute for Risk Assessment, Germany) who generously shared HEV-C positive control as a dried plasmid containing the whole genome of ratHEV strain R63.

The study was supported in part by internal fundings of the Institut Pasteur du Cambodge. The study on hantavirus was a Centers for Research in Emerging Infectious Diseases (CREID) Pilot Research Program 2021 funded by the National Institute of Allergy and Infectious Diseases of the National Institutes of Health under Award Number 1U01AI151378 through Pasteur International Center for Research in Emerging Infectious Diseases (PICREID). The content is solely the responsibility of the authors and does not necessarily represent the official views of the National Institutes of Health.

